# Treatment Heterogeneity of Water, Sanitation, Hygiene, and Nutrition Interventions on Child Growth by Environmental Enteric Dysfunction and Pathogen Status for Young Children in Bangladesh

**DOI:** 10.1101/2024.03.21.24304684

**Authors:** Zachary Butzin-Dozier, Yunwen Ji, Jeremy Coyle, Ivana Malenica, Elizabeth T. Rogawski McQuade, Jessica Anne Grembi, James A. Platts-Mills, Eric R. Houpt, Jay P. Graham, Shahjahan Ali, Md Ziaur Rahman, Mohammad Alauddin, Syeda L. Famida, Salma Akther, Md. Saheen Hossen, Palash Mutsuddi, Abul K. Shoab, Mahbubur Rahman, Md. Ohedul Islam, Rana Miah, Mami Taniuchi, Jie Liu, Sarah Alauddin, Christine P. Stewart, Stephen P. Luby, John M. Colford, Alan E. Hubbard, Andrew N. Mertens, Audrie Lin

**Affiliations:** School of Public Health, University of California, Berkeley, Berkeley, CA USA; Rollins School of Public Health, Emory University, Atlanta, GA USA; Division of Infectious Diseases and Geographic Medicine, Stanford University, Stanford, CA USA; School of Medicine, University of Virginia, Charlottesville, VA, USA; International Centre for Diarrhoeal Disease Research, Bangladesh, Dhaka, Bangladesh; School of Public Health, Qingdao University, Qingdao, China; Wagner College, Staten Island, NY USA; Department of Nutrition, University of California Davis, Davis, CA USA; Department of Microbiology and Environmental Toxicology, University of California, Santa Cruz, Santa Cruz, CA USA

## Abstract

**Background:** Water, sanitation, hygiene (WSH), nutrition (N), and combined (N+WSH) interventions are often implemented by global health organizations, but WSH interventions may insufficiently reduce pathogen exposure, and nutrition interventions may be modified by environmental enteric dysfunction (EED), a condition of increased intestinal permeability and inflammation. This study investigated the heterogeneity of these treatments’ effects based on individual pathogen and EED biomarker status with respect to child linear growth.

**Methods:** We applied cross-validated targeted maximum likelihood estimation and super learner ensemble machine learning to assess the conditional treatment effects in subgroups defined by biomarker and pathogen status. We analyzed treatment (N+WSH, WSH, N, or control) randomly assigned in-utero, child pathogen and EED data at 14 months of age, and child LAZ at 28 months of age. We estimated the difference in mean child length for age Z-score (LAZ) under the treatment rule and the difference in stratified treatment effect (treatment effect difference) comparing children with high versus low pathogen/biomarker status while controlling for baseline covariates.

**Results:** We analyzed data from 1,522 children, who had median LAZ of -1.56. We found that myeloperoxidase (N+WSH treatment effect difference 0.0007 LAZ, WSH treatment effect difference 0.1032 LAZ, N treatment effect difference 0.0037 LAZ) and *Campylobacter* infection (N+WSH treatment effect difference 0.0011 LAZ, WSH difference 0.0119 LAZ, N difference 0.0255 LAZ) were associated with greater effect of all interventions on growth. In other words, children with high myeloperoxidase or *Campylobacter* infection experienced a greater impact of the interventions on growth. We found that a treatment rule that assigned the N+WSH (LAZ difference 0.23, 95% CI (0.05, 0.41)) and WSH (LAZ difference 0.17, 95% CI (0.04, 0.30)) interventions based on EED biomarkers and pathogens increased predicted child growth compared to the randomly allocated intervention.

**Conclusions:** These findings indicate that EED biomarker and pathogen status, particularly *Campylobacter* and myeloperoxidase (a measure of gut inflammation), may be related to impact of N+WSH, WSH, and N interventions on child linear growth.

## Introduction

Approximately 148 million children globally experience linear growth faltering, which may be a consequence of early life undernutrition [1]. Studies have consistently found a positive relationship between child growth and child development, leading investigators to use child linear growth as a proxy for overall development [2,3]. In adulthood, children who experienced early-life growth faltering are more likely to experience low educational attainment and low income, although interventions that improve child growth do not necessarily improve child development (and vice versa) [2–6]. Children of mothers who are stunted have an increased risk of experiencing stunting themselves, which can perpetuate the cycle of poverty [7].

### Water, sanitation, hygiene, and nutrition

Experts in public health and international development have identified water, sanitation, hygiene (WSH), nutrition (N), and combined (N+WSH) programs as potentially effective methods to improve child growth. WSH interventions aim to reduce children’s exposure to pathogens, which can improve nutrient utilization by reducing malabsorption, redirection of nutrients for immune response, and other symptoms associated with infection, while nutrition interventions aim to directly provide nutrient supplementation [8,7]. The United Nations has established universal access to WSH by the year 2030 as a Sustainable Development Goal [9]. Despite the widespread implementation of N and WSH interventions, based on the assumption that these interventions improve child growth, few randomized controlled trials have evaluated the impact of N+WSH, WSH, and N interventions on child growth.

Several observational studies indicated a positive relationship between household WSH interventions and child growth [10]. In contrast to these observational findings, three recent randomized controlled trials in rural populations from Kenya and Bangladesh (the WASH Benefits study) and Zimbabwe (SHINE trial), enrolled pregnant mothers and found that household WSH interventions did not improve child linear growth in a randomized context [7,11–13]. These findings suggested that positive associations between WSH and child growth in observational settings may be due to residual confounding. The null effect of these environmental interventions on growth indicated the possibility that additional sources of growth impairment might exist for children facing extreme poverty. Alternatively, the lack of impact of these interventions may reflect an inability of these household interventions to sufficiently reduce pathogen exposure and environmental enteric dysfunction [11,14,15].

The WASH Benefits study found that nutritional supplementation led to modest improvements in child linear growth compared to control [11]. This is consistent with other randomized controlled trials in low and middle-income countries, which have also found that early nutritional supplementation can improve child growth.[16,17] The combined N+WSH intervention did not provide any additional benefit to child linear growth compared to the nutrition intervention alone [11]. The authors indicated that this small and variable impact of nutrition interventions on child linear growth may be due to contextual underlying factors that influence participants’ ability to respond to and benefit from nutrition interventions [11,18].

### Effect Measure Modification by EED and Pathogens

In addition to finding a null main effect of WSH interventions on growth and modest effects of nutrition on growth (and N+WSH providing no additional benefit compared to nutrition alone), the WASH Benefits study did not detect significant effect modification of interventions by child age, child sex, maternal education, maternal age, child parity, economic factors, or child hunger [11]. Although it should be noted that lack of observed effect measure modification could be due to limited power or the study context [11]. Despite this lack of evidence of interaction, pathogen and environmental enteric dysfunction (EED) biomarker data may provide additional information on which subgroups of children, defined by pathogen or biomarker levels, are amenable or resistant to intervention.

EED is a condition characterized by increased gut permeability, gut barrier disruption, increased gut and systemic inflammation, and is hypothesized to be caused by chronic exposure to pathogens [19,20]. Although clear diagnostic criteria for EED have not been established, several studies have speculated that it could be a key intermediate between poverty and growth impairment for children in low and middle-income countries [19,20]. Observational data and animal models have indicated that *Campylobacter* infection may contribute to EED [21]. Among young children in Bangladesh, small intestine bacterial overgrowth is associated with both intestinal inflammation, a key component of EED, and child growth impairment [22,23]. The WASH Benefits Bangladesh study found that the nutrition intervention was associated with reduction of neopterin at 3 and 14 months of age, and all interventions reduced lactulose and mannitol at 3 and 14 months [24]. At 28 months, contrary to a-priori hypotheses, WSH and nutrition interventions were associated with increased myeloperoxidase, and WSH was associated with increased mannitol absorption [24]. Although these findings at age 3 and 14 months support N+WSH interventions’ ability to reduce some EED biomarkers, the counterintuitive results at 28 months highlight uncertainty regarding the relationship between N+WSH interventions and presumed biomarkers for EED. In the SHINE Trial, investigators found that WSH interventions decreased the number of parasites detected, but did not have a significant effect on bacteria, viruses, or enteropathogens [15].

Investigators of the WASH Benefits study suggested that insufficient reduction of pathogen exposure could explain the null effects of WSH interventions on child linear growth [11].

Investigation of the relationships between N+WSH interventions and enteropathogens at Year 1 (age 14 months) in Bangladesh found that children who received WSH interventions had a lower prevalence and quantity of some individual viruses (norovirus, sapovirus, and adenovirus 40/41) compared to children in the control group, although investigators did not find a significant difference in bacteria, parasites, or stunting-related pathogens between these groups [14].

Furthermore, this study found that 99% of children at Year 1 had at least one enteropathogen [14]. At Year 2 (age 31 months), investigators found that individual sanitation and hygiene interventions were associated with decreased *Giardia* infections and that drinking water and nutrition interventions were not associated with a change in *Giardia* infections [25]. Regarding soil-transmitted helminths, investigators found that the drinking water intervention was associated with reduced hookworm [26]. Lastly, analysis of interventions and fecal contamination found that drinking water and handwashing interventions reduced contamination of water and food, but did not reduce contamination of indirect pathways such as child hands and objects, and that combined WSH interventions provided no additional benefit compared to individual interventions [27]. These cumulative findings indicate that household WSH interventions can reduce child exposure to certain pathogens, although these results highlight heterogenous relationships between interventions and individual pathogens.

### Methodological Utility of Optimal Treatment Regime Analysis

Public health research typically seeks to identify population-level drivers of incidence rates, rather than individual causes of cases [28]. But, methodological advances have enabled the creation of dynamic treatment rules, where susceptible individuals can be targeted for interventions based on individual characteristics or treatment history [29,30]. Despite this focus on optimizing interventions based on individual covariate information, we retain the public health goal of maximizing population-level health outcomes with limited resources [29]. Even if there is a true effect of the intervention on the outcome of interest among certain individuals, a study may fail to detect this relationship if the effect is heterogeneous in the study sample or the subgroup of amenable individuals is small. We can assess the variance of the stratum-specific treatment effect to evaluate treatment heterogeneity [31].

Targeted maximum likelihood estimation is an estimation method that optimizes the bias-variance tradeoff for a specific parameter of interest [32]. This method is efficient when the outcome regression and the treatment mechanism (propensity score) are correctly specified. Due to the doubly-robust property, targeted maximum likelihood estimation returns consistent results as long as either the outcome regression or the treatment mechanism is estimated consistently [32,33].

We can gain additional insight through analysis of optimal individualized treatment effect, where we seek to maximize population outcomes by assigning treatment based on individual characteristics that are associated with the most beneficial treatment effect [34]. Estimation of this optimal individualized treatment effect has gained popularity with the rise in precision health, but much of these efforts have relied on unrealistic parametric assumptions [35–40]. If the parametric model is incorrect (which is often the case), the resulting estimates will be biased for the parameters of interest (e.g., average treatment effect) [34]. Using targeted learning methods, we can estimate the mean outcome under the optimal individualized treatment. The candidate treatment rules are estimated on the same data for which the impact of the rule is also estimated, using a robust cross-validated estimation procedure [34]. One can gain efficiency by constraining the statistical model, so the only restrictions on the data distribution relate to the probability of a participant receiving treatment (which is a fixed, randomized assignment) [34]. The use of cross-validated targeted maximum likelihood estimator for the mean outcome under optimal individualized treatment uses a data-adaptive estimation of the optimal rule, while still providing valid inference on the impact of the estimated optimal treatment without making parametric assumptions.

Using data from the WASH Benefits Bangladesh study, analysis of treatment heterogeneity through estimating subgroup treatment effects and optimal treatment regimens can improve our understanding of child growth in low- and middle-income countries. Despite widespread use of N+WSH interventions, investigators have found mixed evidence regarding these interventions’ impact on child growth [7,11,13]. This study will apply targeted machine learning methods to assess the conditional treatment effect of N+WSH, WSH, and N interventions on child linear growth (child length for age Z score (LAZ)) by pathogen and EED biomarker status and explore rules for the optimal allocation of N+WSH, WSH, and N interventions in resource-constrained settings.

## Methods

### Study design, participants, and interventions

This analysis involves data from a substudy of the WASH Benefits Bangladesh randomized controlled trial. The trial randomized pregnant mothers and their children to receive one of six interventions – water treatment, sanitation, handwashing, nutrition (N), combined water treatment, sanitation and handwashing (WSH) and combined nutrition plus WSH (N+WSH), or control [11]. In a substudy focused on evaluation of EED, investigators assessed additional biomarker data in a subset of children in four of the study arms – N, WSH, N+WSH, and control (with an allocation ratio of 1:1:1:1) [24].

Intervention promoters, who were residents of the study area, visited participants to promote intervention behaviors at the level of the compound (cluster of nearby houses). Each promoter received at least five days of training prior to visiting compounds, and received periodic refresher courses throughout the intervention period. The behavioral components of these interventions included treating drinking water for index households, which included children less than 3 years of age (water), using latrines and child potty in addition to removing animal feces from the compound (sanitation), washing hands with soap before preparing food and after defecating or contacting child feces (hygiene), and practicing age-appropriate nutrition practices from pregnancy up until two years of age and using small-quantity lipid-based nutrient supplements for children six months to two years of age (nutrition) [41]. These promoters used various strategies to promote intervention behaviors. For example, promoters promoted the hygiene intervention (handwashing) by framing it as a nurturing intervention that was facilitated by the handwashing station and soap provided by the intervention [41,42]. Promoters were instructed to visit study compounds at least once per week for the first six months, and then once every two weeks for the following 1.5 years. The intervention hardware and consumables were provided free of charge and replenished by promoters as needed throughout the study period (additional details on interventions can be found in Supplemental Material 1).

Investigators followed the cohort of children for approximately 2.5 years after birth. It was not feasible to retain the geographic matching of the parent trial in this subset due to logistical challenges regarding specimen collection and transportation. The trial was conducted in contiguous rural subdistricts in Gazipur, Mymensingh, Tangail and Kishoreganj districts of Bangladesh. The trial enrolled mothers in their second trimester of pregnancy (additional information on recruitment and eligibility can be found in Supplemental Material 2) [41].

Covariates:

Although randomization of participants led to a balanced distribution of covariates between study arms, this analysis conditioned on post-randomization biomarker values, leading to the possibility of collider stratification bias. In addition to the biomarkers and pathogens included in the treatment rule, our analyses adjusted for child sex, birth order, number of children under 18 years of age in the household, number of individuals in the compound (group of nearby houses), household wall material, household wealth (first principal component of a principal components analysis incorporating household assets), maternal age and height, age in days at urine and stool assessments, month of urine and stool assessments, and age at anthropometry assessment. We considered and tested potential confounders using super learner and cross-validated targeted maximum likelihood estimation. The full list of baseline and time-varying covariates can be found in Supplemental Material 3.

### Biomarkers

#### EED Biomarkers

The EED measures included in this study were fecal alpha-1-antitrypsin, myeloperoxidase, and REG1B, which we measured at median ages 3 months and 14 months. These measures are markers of intestinal permeability (alpha-1-antitrypsin), inflammation (myeloperoxidase), and intestinal repair (REG1B) [43]. We excluded EED biomarkers (neopterin, lactulose, and mannitol) that were associated with the interventions of interest in a previous analysis of this sample and therefore were potential mediators of the exposure-outcome relationship [24].

To reduce inter-laboratory variation, all fecal samples were assayed by the same research team member at the International Centre for Diarrhoeal Disease Research, Bangladesh (icddr,b) laboratory. Laboratory methods are included in Supplemental Material 4 and were published previously [14,24].

#### Pathogens

We included six pathogens in our final analysis: *Campylobacter jejuni/coli*, enteroaggregative *Escherichia coli* (EAEC), any enterotoxigenic *E. coli* (ETEC), atypical enteropathogenic *E. coli* (aEPEC), any enteropathogenic *E. coli* (EPEC, including both typical or atypical), and *Campylobacter* spp. (which includes *C. jejuni, C. coli,* and other *Campylobacter* species).

Relative concentrations of pathogens were assessed at 14 months in feces using quantitative polymerase chain reaction (qPCR) via TaqMan array card [14,44,45]. We excluded three pathogens (norovirus, sapovirus, and adenovirus 40/41) that were reduced by the interventions of interest in a previous analysis of this sample and therefore were potential mediators of the exposure-outcome relationship [14]. We excluded an additional 25 pathogens due to high missingness or near-zero variance. We quantified pathogens via quantification cycle, where one unit represented twice the pathogen quantity, and the analytical limit of detection was at quantification cycle 35 [46]. We standardized these measures using the efficiency of per-sample extraction/amplification. The full list of pathogens is included in Supplemental Material 5.

A single infection event is unlikely to elicit growth impairment in itself, but repeated exposure to pathogens and chronic disruptions such as EED are associated with delayed growth [47–49].

This analysis assumes that the detection of pathogens and EED biomarkers at 14 months is a proxy for chronic exposure to these factors throughout early childhood.

#### Analyzed biomarkers

Our treatment rule included the pathogens *Campylobacter jejuni/coli*, EAEC, ETEC, aEPEC, EPEC, and *Campylobacter* spp., as well as EED biomarkers regenerating gene 1β (REG1B), myeloperoxidase, and alpha-1-antitrypsin.

### Outcomes

The growth outcome was length for age Z-score (LAZ) assessed at Year 2 (median age 28 months). Following standard protocols for anthropometric outcomes measurement [50,51], pairs of trained anthropometrists measured child growth (accurate to 0.1 cm) in triplicate to calculate median growth using 2006 WHO child growth standards [11]. We measured recumbent length when child was age < 24 months. We measured standing height when child was age > 24 months.

### Analyses

These analyses assessed the conditional average treatment effect (CATE) and mean under the optimal individualized treatment regime using a targeted learning approach [34]. A static treatment approach as used in the WASH Benefits primary analysis, in which treatment is randomly assigned, aims to assess the average effect of the interventions in the entire study population (i.e. interventions are not targeted based on individual covariate information) [11,34]. In contrast, an optimal treatment regime analysis assesses the impact of the intervention given (or, conditional on) individual covariate status [11,34]. In these analyses, the individual covariate information was child pathogen and EED biomarker status.

We used cross-validated targeted maximum likelihood estimation, which we fit using Super Learner ensemble machine learning in order to estimate the optimal individualized treatment regime and the outcome as the mean under the optimal individualized treatment [31]. First, we estimated the outcome regression function and propensity score (treatment mechanism) using Super Learner. Super Learner is an ensemble machine learning approach that creates a convex combination of candidate algorithms in order to maximize model fit [52,53]. Super Learner is grounded in statistical optimality theory, and guarantees that it will perform at least as well as the best candidate algorithm with a sufficiently large sample size. In our learner list for the treatment mechanism, we included the least absolute shrinkage and selection operator (LASSO) penalized regressions, random forests, the simple mean, and generalized linear models, and we used non-negative least squares to construct the final ensemble (the meta-learner) [34].

Next, we used the doubly-robust augmented inverse probability weighting to transform the outcome to a random variable that has as the CATE (i.e., the treatment effect specific to each individual’s set of covariates) as its mean and regressed this transformed outcome to assess treatment heterogeneity using targeted maximum likelihood estimation via the R package “tmle3mopttx” [54]. Targeted maximum likelihood estimation reduces bias and yields an interpretable measure of association (in this case, the average treatment effect) [34,55–58].

Specifically, we estimated the function of the individualized outcome by regressing this contrast on biomarker status using Super Learner with a non-negative least squares loss function based on the Lawson-Hanson algorithm. As these analyses assess the impact of the randomized intervention (the treatment mechanism), the doubly-robust nature of this estimator will ensure asymptotically consistent estimation of the CATE even if the outcome regression is not consistently estimated [34].

Finally, we use the estimate of the CATE function to derive an optimal individualized treatment rule where we would treat a maximum of 50% of individuals with the greatest CATE. Though providing optimal treatment to all children is desirable, in a resource constrained setting, one might also be interested to limit the intervention to the children most likely to benefit from the intervention (i.e., have the greatest CATE). In order to assess the impact of the individualized treatment regime in resource-constrained settings (i.e., preventing all children from being allocated to intervention), we restricted the maximum allocation to treatment in each binary (treatment to control) contrast to be no more than 50%, which is approximately equivalent to the original trial’s allocation ratio (1:1:1:1). If less than 50% of individuals in a single binary (treatment to control) contrast have a positive CATE (beneficial effect of treatment), then the optimal treatment rule will assign all individuals with a positive CATE to intervention. If more than 50% of individuals in a single contrast have a positive CATE, the optimal treatment rule will only assign the 50% of individuals with the greatest CATE to intervention.

In order to assess the role of each biomarker or pathogen in the optimal treatment rule, we evaluated Pearson’s correlation between each of these covariates and the CATE. In order to contextualize the magnitude of these relationships, we estimated the difference in subgroup treatment effect between children with high (detection for pathogens, above median for EED biomarkers) versus low (non-detection for pathogens, below median for EED biomarkers) pathogen and EED biomarker status. While the optimal treatment rule flexibly incorporated continuous values of these biomarkers, we used binary transformations of these values to assess variable importance in order to improve interpretability. Furthermore, while the optimal treatment rule assessed the combined role of these biomarkers, our assessments of variable importance assessed each biomarker individually to improve interpretability. The difference between these two subgroup effects is hereafter referred to as “treatment effect difference.”

#### Covariate screening

We screened all covariates for missingness, excluding all covariates with missingness greater than 30% and median-imputing all other missing covariate data. We only included observations for which the primary outcome, LAZ at 28 months, was observed. We also excluded variables with near zero variance, which we defined as covariates with a frequency ratio (ratio of most frequent value to second most frequent value) greater than 2 and a percent of unique values less than 20%, using the R package “caret” (version 6.0-92) [59]. The analysis plan was publicly pre-registered on Open Science Framework, and all data and analysis scripts are publicly available (https://osf.io/cg8dv/). EED markers assessed at 3 months were excluded due to high missingness (>30%). The full list of excluded covariates and reasons for exclusion are defined in Supplemental Material 6.

#### Ethics

The primary caregiver of each child provided written informed consent prior to enrollment. Human subjects protection committees at icddr,b, the University of California, Berkeley, and Stanford University approved the study protocols. The parent trial was registered at ClinicalTrials.gov (NCT01590095) and a safety monitoring committee convened by icddr,b oversaw the study.

## Results

We analyzed data from 1,522 children, and our analytic sample had a median LAZ of -1.56 at Year 2 (median age 28 months; Table 1).

**Table 1.**
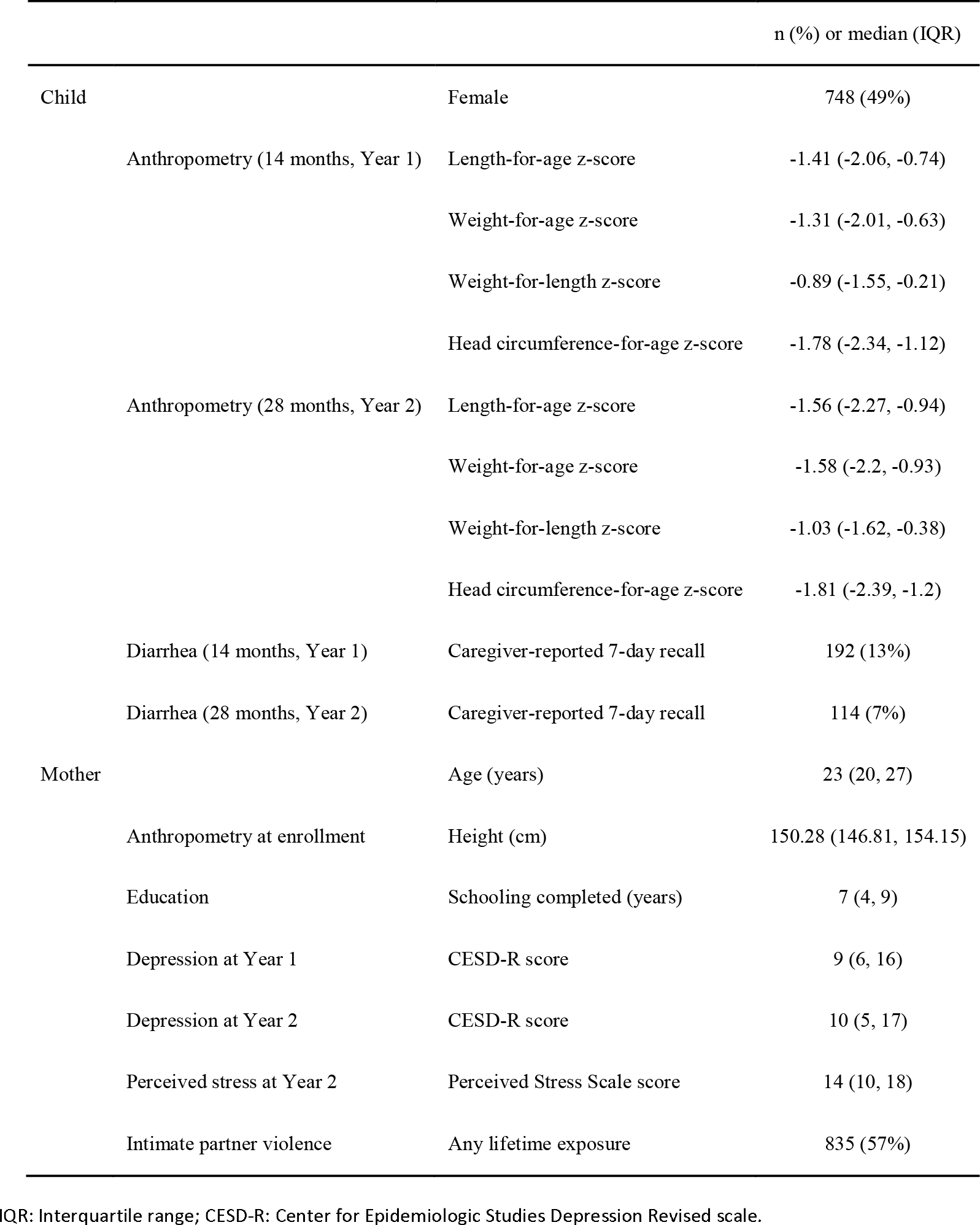
Descriptive statistics of sample population.

### Relationships between pathogens or biomarkers and the conditional average treatment effect

In order to identify subgroups of children (based on EED biomarker and pathogen values) with the largest treatment effect, we analyzed the treatment effect comparing children with high (detection for pathogens, above median for EED biomarkers) versus low (non-detection for pathogens, below median for EED biomarkers) values of each pathogen or biomarker.

We found that the following covariates were associated with a greater impact of N+WSH intervention on growth under the optimal treatment rule: ETEC (correlation 0.45, treatment effect difference (comparing the treatment among children with high ETEC to children with low ETEC) 0.0019 LAZ)*, Campylobacter jejuni/coli* (correlation 0.37, treatment effect difference 0.0016 LAZ), *Campylobacter* spp. (correlation 0.33, treatment effect difference 0.0011 LAZ), REG1B (correlation 0.20, treatment effect difference 0.0005 LAZ), and myeloperoxidase (correlation 0.15, treatment effect difference 0.0007 LAZ) (Table 2). The following covariates were associated with a lower impact of N+WSH intervention: aEPEC (correlation -0.41, treatment effect difference -0.0018 LAZ), EAEC (correlation -0.39, treatment effect difference -0.0015 LAZ), alpha-1-antitrypsin (correlation -0.38, treatment effect difference -0.0013 LAZ), and EPEC (correlation -0.22, treatment effect difference -0.0009 LAZ).

**Table 2.**
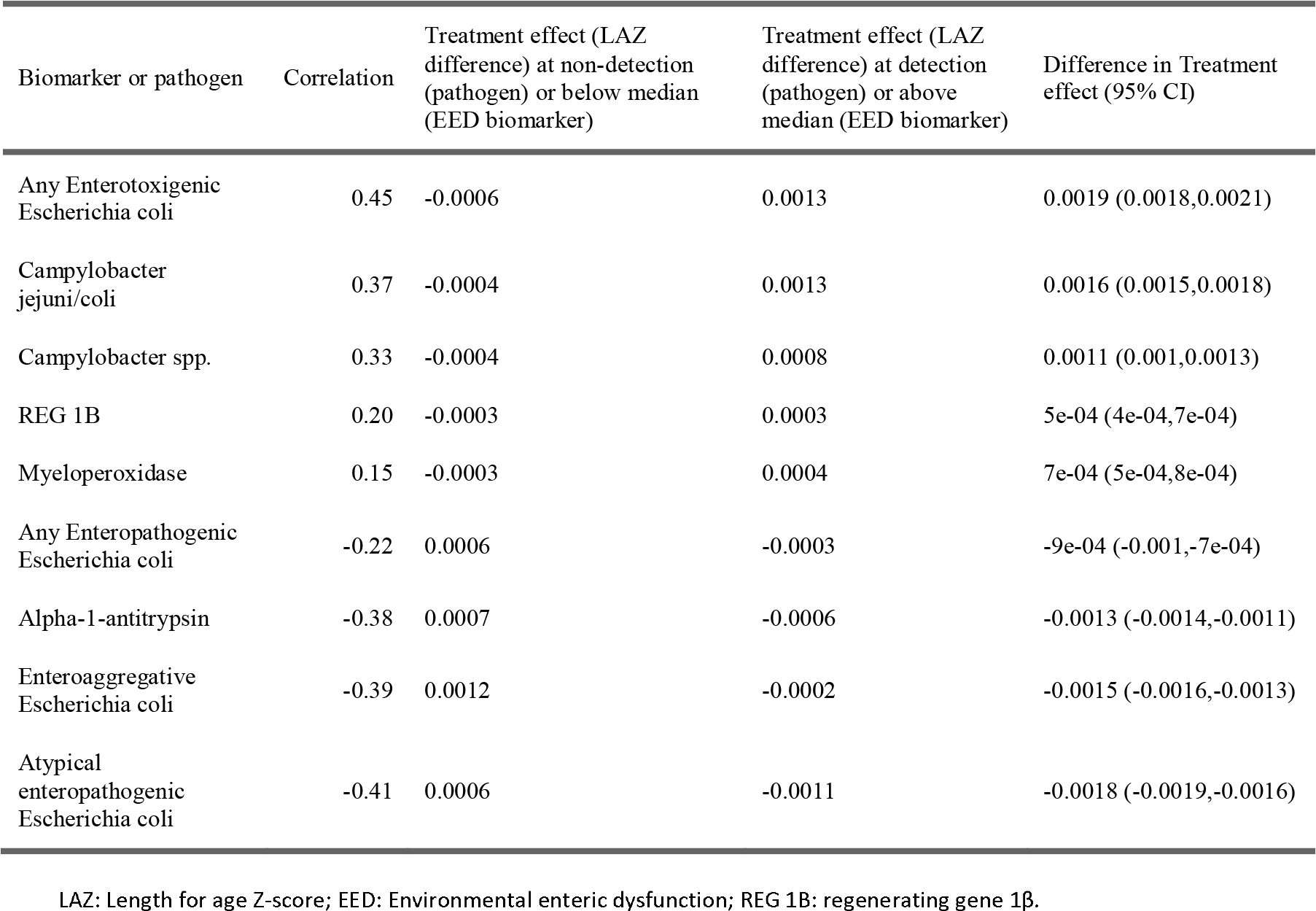
Biomarker and Pathogen Correlation with NWSH Conditional Average Treatment effect.

The following EED biomarkers and pathogens were associated with greater WSH impact on growth under the optimal treatment rule: myeloperoxidase (correlation 1.00, treatment effect difference 0.1032 LAZ), alpha-1-antitrypsin (correlation 0.26, treatment effect difference 0.0259 LAZ), REG1B (correlation 0.17, treatment effect difference 0.0105 LAZ), *Campylobacter jejuni/coli* (correlation 0.15, treatment effect difference 0.0143), *Campylobacter* spp. (correlation 0.13, treatment effect difference 0.0119 LAZ), EPEC (correlation 0.11, treatment effect difference 0.014 LAZ), and aEPEC (correlation 0.08, treatment effect difference 0.0099 LAZ) (Table 3). No EED biomarkers or pathogens were associated with lower WSH treatment effect.

**Table 3.**
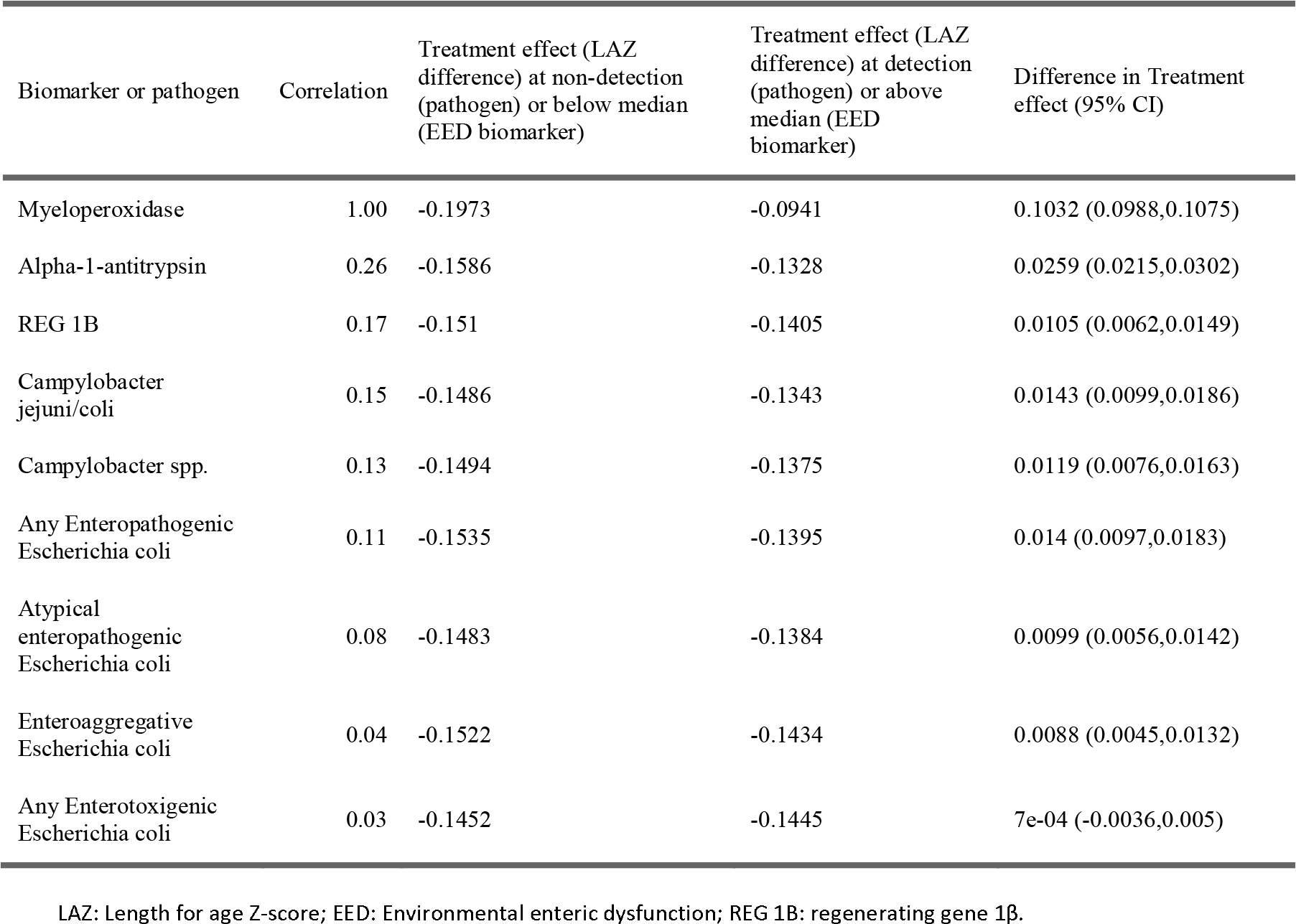
Biomarker and Pathogen Correlation with Conditional Average WSH Treatment effect.

The following EED biomarkers and pathogens were associated with greater impact of N on growth under the optimal treatment rule: *Campylobacter* spp. (correlation 0.17, treatment effect difference 0.0255 LAZ), *Campylobacter jejuni/coli* (correlation 0.15, treatment effect difference 0.0269 LAZ), myeloperoxidase (correlation 0.06, treatment effect difference 0.0037 LAZ), and ETEC (correlation 0.05, treatment effect difference 0.0098 LAZ) (Table 4). EAEC (correlation - 0.07, treatment effect difference -0.0181 LAZ) was associated with a lower impact of N intervention.

**Table 4.**
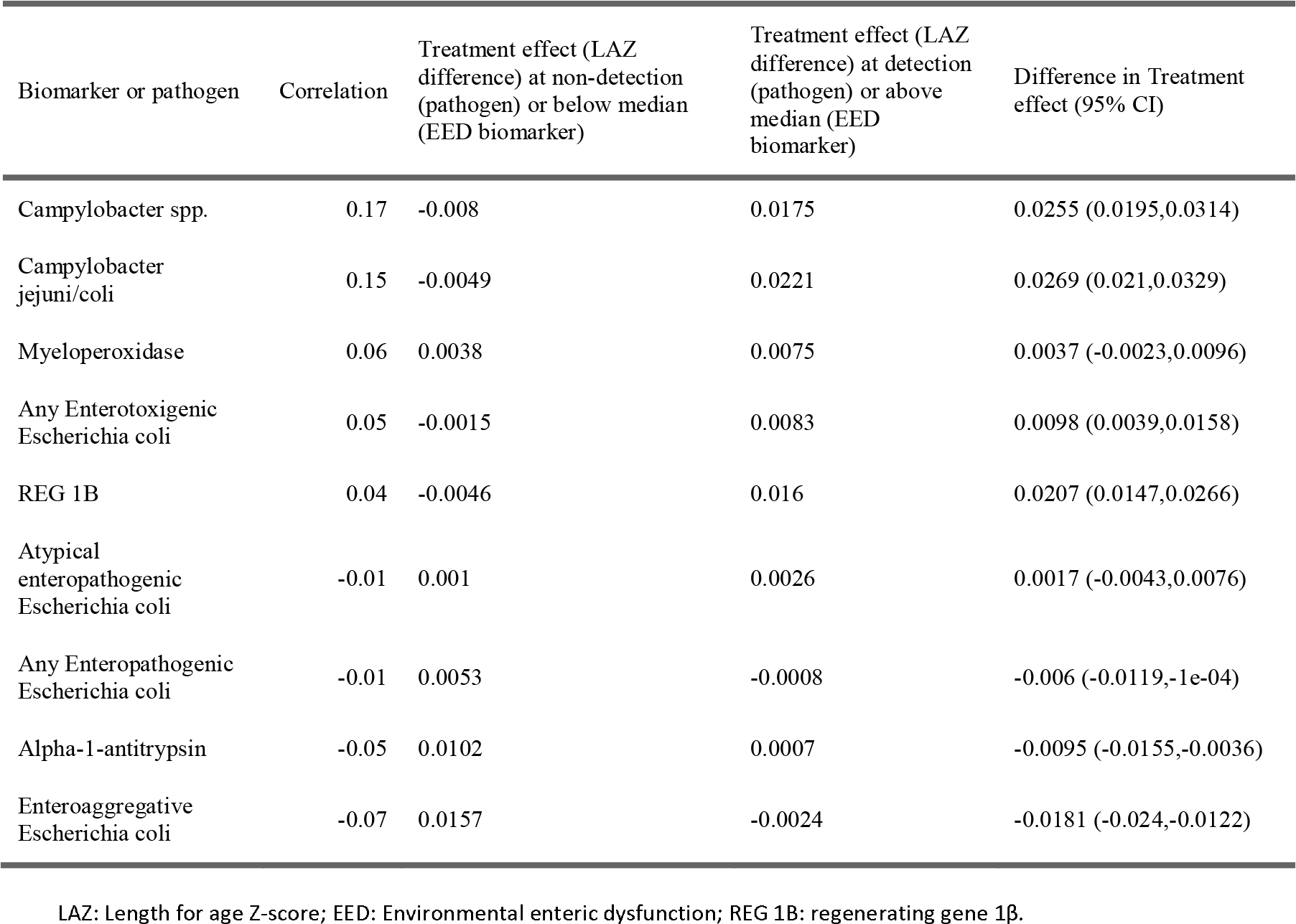
Biomarker and Pathogen Correlation with Nutrition Conditional Average Treatment Effect.

### Treatment allocation and predicted child growth

When comparing the combined N+WSH (mean LAZ -1.62) and control (mean LAZ -1.54) arms (*n* = 756), an optimal treatment allocation assigned 331 children to N+WSH and 425 children to control (Table 5). The optimal treatment rule predicted greater child growth than the observed randomized intervention (observed LAZ -1.58 vs. optimal LAZ -1.35; optimal vs. observed growth difference 0.23 LAZ, 95% CI (0.05, 0.41)).

**Table 5.** Average child growth given optimized vs randomized treatment.

| Study arms | n | Observed growth in treatment arm | Observed growth in control arm | Optimal allocation ratio (treatment: control) | Overall observed child growth | Optimized child growth | Predicted growth difference |
| --- | --- | --- | --- | --- | --- | --- | --- |
| N+WSH vs. control | 756 | -1.62 | -1.54 | (331:425) | -1.58 | -1.35 ( -1.53 , -1.17 ) | 0.23 ( 0.05 , 0.41 ) |
| WSH vs. control | 752 | -1.69 | -1.54 | (9:743) | -1.62 | -1.45 ( -1.58 , -1.32 ) | 0.17 ( 0.04 , 0.3 ) |
| Nutrition vs. control | 726 | -1.53 | -1.54 | (317:409) | -1.53 | -1.47 ( -1.62 , -1.31 ) | 0.07 ( -0.09 , 0.22 ) |

N+WSH: Combined nutrition, water, sanitation, and hygiene intervention; WSH: Water, sanitation, and hygiene intervention.

In the contrast of WSH (mean LAZ -1.69) and control (mean LAZ -1.54) arms (*n* = 752), the optimal treatment rule assigned 9 children to receive WSH interventions and 743 children to receive control. The optimal treatment rule had greater predicted child growth than the observed randomized, static intervention (observed LAZ -1.62 vs. optimal LAZ -1.45; optimal vs. observed growth difference 0.17 LAZ, 95% CI (0.04, 0.3)).

After comparing the nutrition (mean LAZ -1.53) and control (mean LAZ -1.54) arms (*n* = 726), the optimal treatment rule assigned 317 children to receive the intervention and 409 children to be in the control group. The optimal treatment rule did not have significantly greater child growth compared to the observed randomized intervention (observed LAZ -1.53 vs. optimal LAZ -1.47; optimal vs. observed growth difference 0.07 LAZ, 95% CI (-0.09, 0.22).

### Post-hoc analysis

*Campylobacter* spp. and myeloperoxidase were associated with a greater treatment effect across all three interventions (Supplemental Materials 7-12). We conducted an exploratory evaluation of the combined impact of *Campylobacter* infection (any detection) and high myeloperoxidase (above median concentration) on the conditional treatment effect under the optimal treatment rule (Table 6). The difference in treatment effect, comparing those with both *Campylobacter* spp. infection and high myeloperoxidase to those with no *Campylobacter* spp. detection and below median myeloperoxidase, was 0.039 LAZ for N+WSH, 0.106 LAZ for WSH, and 0.022 LAZ for N.

**Table 6.** Conditional average treatment effect given levels of both Campylobacter and myeloperoxidase at 14 months.

| Treatment arm | Treatment effect (LAZ difference) given Campylobacter non-detection and below median myeloperoxidase | Treatment effect (LAZ difference) given Campylobacter detection and above median myeloperoxidase | Difference in treatment effect (LAZ difference) | Difference 95% CI |
| --- | --- | --- | --- | --- |
| N+WSH | -0.0007 | 0.0011 | 0.0018 | 0.0017 , 0.002 |
| WSH | -0.1981 | -0.0917 | 0.1064 | 0.1021 , 0.1107 |
| Nutrition | -0.0029 | 0.0187 | 0.0216 | 0.0157 , 0.0276 |

N+WSH: Combined nutrition, water, sanitation, and hygiene intervention; WSH: Water, sanitation, and hygiene intervention; LAZ: Length for age Z-score

## Discussion

Across all three interventions, myeloperoxidase, an EED biomarker of gut inflammation, and *Campylobacter* were associated with a greater treatment effect [43,60]. In other words, children with the greatest burden of *Campylobacter* infection and myeloperoxidase experienced the greatest benefit from the interventions, although the magnitude of these differences in treatment effects was typically small and not clinically significant. There was a greater N+WSH and WSH treatment effect among those with both *Campylobacter* infection and high myeloperoxidase than those with either factor alone. The correlation of both *Campylobacter* and myeloperoxidase biomarkers with the treatment effect indicates that these factors, implicated as a cause (*Campylobacter*) and a marker (myeloperoxidase) of EED, supports that EED may play a role in the interventions’ impact on growth [47]. These results are consistent with previous findings that young children with *Campylobacter* infection may face increased risk of growth impairment and are therefore a high-need group for intervention. A multi-site birth cohort study (MAL-ED) found that *Campylobacter* infection was highly prevalent and was associated with decreased child growth in the first two years of life [61,48]. *Campylobacter* infections are endemic in settings where poultry is raised near the household (which is common in low and middle income countries), and even asymptomatic infection is negatively associated with child growth [48,62,63]. *Campylobacter* alters the gut microbiota composition, disrupts the intestinal barrier, and can elicit chronic intestinal inflammation [64–69]. Across eight study sites in low-resource settings, MAL-ED found that breastfeeding, lack of access to WSH, and targeted antibiotic treatment were associated with *Campylobacter* infection [48]. In addition, these investigators found that *Campylobacter* infection was associated with increased intestinal permeability, intestinal inflammation, and systemic inflammation, which are key components of EED [48].

We found that myeloperoxidase, an EED marker of gut inflammation, was associated with a greater impact of N+WSH, WSH, and N interventions on child growth. That is to say, children with higher intestinal inflammation were most protected by the interventions. This is likely indicative of children whose household environments were the most contaminated and interventions reduced but did not eliminate environmental exposures, as gut inflammation remained high despite continued intervention delivery. Previous meta-analyses have found that inflammation and WSH conditions modify the effects of nutrient supplementation on micronutrient status and anemia [16,70]. Regarding WSH, our findings were consistent with MAL-ED’s findings that EED and inflammation likely mediated the relationship between infection and growth faltering [47]. In addition, MAL-ED investigators found that myeloperoxidase was associated with pathogen infection, and more specifically, that *Campylobacter* and myeloperoxidase were positively associated across all eight study sites [71].

After comparing both WSH and combined N+WSH interventions to control, we found that an optimal treatment rule selected via cross-validation and based on EED and pathogen status led to greater expected mean child growth than the observed, randomized intervention. This indicates that pathogen and EED biomarker status may define, in part, which children are responsive to WSH and N+WSH interventions.

There was a large disparity between the individual biomarker treatment effect differences (very small) and the overall shift in growth under the optimal treatment regime (moderate). The optimal treatment regime takes all of the factors into account, while the treatment effect difference only looks at biomarkers one at a time. Next, the treatment effect difference dichotomizes all of the biomarkers, while the optimal treatment regime incorporates their continuous values in whatever way is most informative to the optimal treatment regime. The disparity between these values highlights how a flexible nonparametric approach such as Targeted Machine Learning can outperform parametric specification of subgroups. These findings highlight the potential for targeted learning methods to identify and explore treatment heterogeneity within a study and for optimal treatment regime analysis to estimate the effects of targeting treatments to children who would benefit the most when resource constraints prevent intervening on all children.

These findings provide empirical support for the hypothesis that pathogen exposure and EED biomarkers are associated with growth faltering. Within rural Bangladesh, these effects were small, but they provide support for a biological mechanism.

### Strengths

The rich data source of the WASH Benefits Bangladesh EED substudy is a major strength of this analysis. This data source included *in utero* randomized interventions that were continued for two years after birth and robust collection of enrollment covariates, EED biomarkers, pathogens, and growth outcomes across multiple timepoints. Furthermore, the statistical methods applied here allow us to flexibly assess relationships between multiple covariates, exposures, and outcomes while making minimal parametric assumptions.

The analysis methods are a second major strength of this study. We used targeted maximum likelihood estimation, which is maximally efficient in finite samples and doubly-robust [32,33]. Assessment of optimal individualized treatment effects allows us to evaluate the relationships between pathogen exposure, EED, and intervention effects without making parametric assumptions [34–40]. Given the complex relationships between these biomarker and pathogen data, these targeted learning methods allow flexible modeling of complex relationships without requiring parametric assumptions regarding relationships between interventions, biomarkers, pathogens, and child growth that would inevitably be violated.

### Limitations

One limitation of this study arises from using post-intervention biomarkers, as no baseline EED biomarkers or pathogens were measured because infants were *in utero* at the time of randomization. Conditioning on these post-intervention nodes potentially introduces confounding and bias. We accounted for this possible confounding by adjusting for additional baseline covariate information related to family health and socioeconomic status and by excluding pathogens and EED biomarkers that were associated with the interventions in previous analyses of this sample (i.e., potential mediators or colliders), although residual confounding or bias may be present. However, the identification of these relationships remains useful for generating hypotheses about the causes of N+WSH, WSH, and N treatment heterogeneity. In the future, we hope to analyze biological samples that were collected from these children at a younger age (4-8 months) in order further evaluate these relationships. Furthermore, future studies should evaluate these relationships by assessing biomarker and pathogen status prior to randomization. In addition, the limited external validity of randomized controlled trials is a limitation of these findings, as these conclusions may not be generalizable to populations outside these communities. The external validity of these findings is also limited due to the trial context, in which participating households received extensive follow-up and monitoring and may not reflect the experience of these interventions in the target population’s context.

The small or null overall effects of the study interventions is another limitation. In the presence of a null overall effect, in order to detect subpopulations that have a significant effect, there must be equivalent populations with a deleterious effect or much larger populations with a null effect. In contrast, optimal treatment regime analysis in a population with a greater treatment effect will have much greater power to detect subpopulations of interest. In other words, optimal treatment regime analyses have the most power to detect treatment heterogeneity in study settings where there is a large overall treatment effect, but this sample had a null overall treatment effect.

Furthermore, the subsample analyzed here did not retain the same growth characteristics as the total trial population. While the trial reported that N and N+WSH interventions led to a modest improvement in growth [11], these effects were not seen for this subsample. This may be the reason that more than half of the children were assigned to control rather than the interventions in the optimal treatment regime, which should be taken as a finite sample limitation of a trial with null effects on children within the small substudy. Follow up evaluation of these relationships in a separate population may provide insight on the replicability of these findings.

### Future directions

These findings support the application and evaluation of interventions that aim to reduce exposure to pathogens, such as *Campylobacter*, as well as interventions that seek to reduce markers of inflammation, such as myeloperoxidase. Evaluations of these interventions should evaluate their direct impacts on these biomarkers as well as their indirect impacts on child growth.

We found that the interventions had the greatest effect in children with a high burden of pathogens and EED biomarkers. Future evaluations that consistently identify biomarkers associated with lower treatment effect (i.e., resistance to treatment) could indicate the need for co-interventions. For example, certain types of persistent bacterial infection (e.g., *Mycobacterium tuberculosis* or *Salmonella typhi*) may not be responsive to WSH interventions, and may require additional medical intervention [72–74]. In these cases, co-interventions, such as antibiotic treatment, may supplement interventions in order to ameliorate these conditions and improve N+WSH, WSH, or N intervention effectiveness [72].

We focused our interpretation on *Campylobacter* and myeloperoxidase, which demonstrated consistent correlations (in terms of direction) with the CATE across interventions. Our analysis of individual biomarkers’ and pathogens’ correlations with the conditional treatment effect provided some evidence of effect heterogeneity being associated with factors beyond *Campylobacter* and myeloperoxidase, although the lack of consistency of these observations across similar interventions (e.g., N+WSH versus WSH) led us to believe that these relationships may be spurious. On the other hand, it is plausible that these unique correlations across similar biomarkers point to unique actions of related covariates or unique mechanisms of combined versus individual interventions, respectively. Future studies could incorporate cluster analysis methods to assess the combined role of related biomarkers and pathogens on treatment effectiveness.

## Conclusion

The cumulative results here indicate that EED and pathogens may be related to N+WSH, WSH, and N interventions’ impact on child growth. In particular, we found that *Campylobacter* infection and high myeloperoxidase were associated with a greater effect of N+WSH (treatment effect difference 0.039 LAZ), WSH (treatment effect difference 0.106 LAZ), and N (treatment effect difference 0.022 LAZ) interventions on child LAZ at 28 months. These findings are consistent with the observational results of the MAL-ED study [47,48,75]. This information regarding the relationships between pathogens, EED biomarkers, and treatment effectiveness highlights biological mechanisms that may indicate an individual’s ability to respond to N+WSH, WSH, and N interventions. These results may help distinguish what defines a responsive versus nonresponsive individual to these interventions and should motivate future etiological research that seeks to estimate the causal impact of EED and pathogen burden on intervention effectiveness.

## Data Availability

All data and analysis scripts are publicly available via

https://osf.io/cg8dv/

## Acknowledgments

We thank the families who participated in the WASH Benefits study and the incredible icddr,b staff for their valuable contributions. This work was supported by Global Development grants [OPPGD759 and OPP1165144] from the Bill & Melinda Gates Foundation to the University of California, Berkeley and by the National Institute of Allergy and Infectious Diseases of the National Institutes of Health [grant number K01AI136885 to AL]. icddr,b is grateful to the Governments of Bangladesh and Canada for providing core/unrestricted support.

## Disclaimer

The content is solely the responsibility of the authors and does not necessarily represent the official views of the National Institutes of Health.

## References

1. United Nations Children’s Fund (UNICEF), World Health Organization (WHO), International Bank for Reconstruction and Development/The World Bank. Levels and trends in child malnutrition: UNICEF / WHO / World Bank Group Joint Child Malnutrition Estimates: Key findings of the 2023 edition. UNICEF and WHO; 2023. Available: https://www.who.int/data/gho/data/themes/topics/joint-child-malnutrition-estimates-unicef-who-wb

2. Grantham-McGregor S, Cheung YB, Cueto S, Glewwe P, Richter L, Strupp B. Developmental potential in the first 5 years for children in developing countries. The Lancet. 2007;369: 60–70. doi:10.1016/S0140-6736(07)60032-4

3. Sudfeld CR, McCoy DC, Danaei G, Fink G, Ezzati M, Andrews KG, et al. Linear growth and child development in low- and middle-income countries: a meta-analysis. Pediatrics. 2015;135: e1266–1275. doi:10.1542/peds.2014-3111

4. Prado EL. Children Staying Smaller but Growing Smarter Beyond the First 1000 Days. J Nutr. 2021;151: 1684–1685. doi:10.1093/jn/nxab136

5. Prado EL, Larson LM, Cox K, Bettencourt K, Kubes JN, Shankar AH. Do effects of early life interventions on linear growth correspond to effects on neurobehavioural development? A systematic review and meta-analysis. The Lancet Global Health. 2019;7: e1398–e1413. doi:10.1016/S2214-109X(19)30361-4

6. Leroy JL, Frongillo EA. Perspective: What Does Stunting Really Mean? A Critical Review of the Evidence. Adv Nutr. 2019;10: 196–204. doi:10.1093/advances/nmy101

7. Pickering AJ, Null C, Winch PJ, Mangwadu G, Arnold BF, Prendergast AJ, et al. The WASH Benefits and SHINE trials: interpretation of WASH intervention effects on linear growth and diarrhoea. The Lancet Global Health. 2019;7: e1139–e1146. doi:10.1016/S2214-109X(19)30268-2

8. Katona P, Katona-Apte J. The Interaction between Nutrition and Infection. Clinical Infectious Diseases. 2008;46: 1582–1588. doi:10.1086/587658

9. Hutton G, Chase C. Water Supply, Sanitation, and Hygiene. 3rd ed. In: Mock CN, Nugent R, Kobusingye O, Smith KR, editors. Injury Prevention and Environmental Health. 3rd ed. Washington (DC): The International Bank for Reconstruction and Development / The World Bank; 2017. doi:10.1596/978-1-4648-0522-6_ch9

10. Cumming O, Cairncross S. Can water, sanitation and hygiene help eliminate stunting? Current evidence and policy implications. Matern Child Nutr. 2016;12 Suppl 1: 91–105. doi:10.1111/mcn.12258

11. Luby SP, Rahman M, Arnold BF, Unicomb L, Ashraf S, Winch PJ, et al. Effects of water quality, sanitation, handwashing, and nutritional interventions on diarrhoea and child growth in rural Bangladesh: a cluster randomised controlled trial. Lancet Glob Health. 2018;6: e302–e315. doi:10.1016/s2214-109x(17)30490-4

12. Gough EK, Moulton LH, Mutasa K, Ntozini R, Stoltzfus RJ, Majo FD, et al. Effects of improved water, sanitation, and hygiene and improved complementary feeding on environmental enteric dysfunction in children in rural Zimbabwe: A cluster-randomized controlled trial. PLoS Negl Trop Dis. 2020;14: e0007963. doi:10.1371/journal.pntd.0007963

13. Null C, Stewart CP, Pickering AJ, Dentz HN, Arnold BF, Arnold CD, et al. Effects of water quality, sanitation, handwashing, and nutritional interventions on diarrhoea and child growth in rural Kenya: a cluster-randomised controlled trial. Lancet Glob Health. 2018;6: e316–e329. doi:10.1016/S2214-109X(18)30005-6

14. Grembi JA, Lin A, Karim MA, Islam MO, Miah R, Arnold BF, et al. Effect of water, sanitation, handwashing and nutrition interventions on enteropathogens in children 14 months old: a cluster-randomized controlled trial in rural Bangladesh. J Infect Dis. 2020. doi:10.1093/infdis/jiaa549

15. Rogawski McQuade ET, Platts-Mills JA, Gratz J, Zhang J, Moulton LH, Mutasa K, et al. Impact of Water Quality, Sanitation, Handwashing, and Nutritional Interventions on Enteric Infections in Rural Zimbabwe: The Sanitation Hygiene Infant Nutrition Efficacy (SHINE) Trial. J Infect Dis. 2020;221: 1379–1386. doi:10.1093/infdis/jiz179

16. Dewey KG, Wessells KR, Arnold CD, Prado EL, Abbeddou S, Adu-Afarwuah S, et al. Characteristics that modify the effect of small-quantity lipid-based nutrient supplementation on child growth: an individual participant data meta-analysis of randomized controlled trials. Am J Clin Nutr. 2021;114: 15S–42S. doi:10.1093/ajcn/nqab278

17. Dewey KG, Arnold CD, Wessells KR, Prado EL, Abbeddou S, Adu-Afarwuah S, et al. Preventive small-quantity lipid-based nutrient supplements reduce severe wasting and severe stunting among young children: an individual participant data meta-analysis of randomized controlled trials. Am J Clin Nutr. 2022;116: 1314–1333. doi:10.1093/ajcn/nqac232

18. Dewey KG, Stewart CP, Wessells KR, Prado EL, Arnold CD. Small-quantity lipid-based nutrient supplements for the prevention of child malnutrition and promotion of healthy development: overview of individual participant data meta-analysis and programmatic implications. Am J Clin Nutr. 2021;114: 3S–14S. doi:10.1093/ajcn/nqab279

19. Watanabe K, Petri WAJ. Environmental Enteropathy: Elusive but Significant Subclinical Abnormalities in Developing Countries. EBioMedicine. 2016;10: 25–32. doi:10.1016/j.ebiom.2016.07.030

20. Ali A, Iqbal NT, Sadiq K. Environmental enteropathy. Curr Opin Gastroenterol. 2016;32: 12–17. doi:10.1097/MOG.0000000000000226

21. Giallourou N, Medlock GL, Bolick DT, Medeiros PH, Ledwaba SE, Kolling GL, et al. A novel mouse model of Campylobacter jejuni enteropathy and diarrhea. PLOS Pathogens. 2018;14: e1007083. doi:10.1371/journal.ppat.1007083

22. Donowitz Jeffrey R., Haque Rashidul, Kirkpatrick Beth D., Alam Masud, Lu Miao, Kabir Mamun, et al. Small Intestine Bacterial Overgrowth and Environmental Enteropathy in Bangladeshi Children. mBio. 7: e02102–15. doi:10.1128/mBio.02102-15

23. Donowitz JR, Pu Z, Lin Y, Alam M, Ferdous T, Shama T, et al. Small Intestine Bacterial Overgrowth in Bangladeshi Infants Is Associated With Growth Stunting in a Longitudinal Cohort. Official journal of the American College of Gastroenterology | ACG. 2022;117. Available: https://journals.lww.com/ajg/Fulltext/2022/01000/Small_Intestine_Bacterial_Overgrowth_i n.26.aspx

24. Lin A, Ali S, Arnold BF, Rahman MZ, Alauddin M, Grembi J, et al. Effects of Water, Sanitation, Handwashing, and Nutritional Interventions on Environmental Enteric Dysfunction in Young Children: A Cluster-randomized, Controlled Trial in Rural Bangladesh. Clinical Infectious Diseases. 2020;70: 738–747. doi:10.1093/cid/ciz291

25. Lin A, Ercumen A, Benjamin-Chung J, Arnold BF, Das S, Haque R, et al. Effects of Water, Sanitation, Handwashing, and Nutritional Interventions on Child Enteric Protozoan Infections in Rural Bangladesh: A Cluster-Randomized Controlled Trial. Clin Infect Dis. 2018;67: 1515–1522. doi:10.1093/cid/ciy320

26. Ercumen A, Benjamin-Chung J, Arnold BF, Lin A, Hubbard AE, Stewart C, et al. Effects of water, sanitation, handwashing and nutritional interventions on soil-transmitted helminth infections in young children: A cluster-randomized controlled trial in rural Bangladesh. PLOS Neglected Tropical Diseases. 2019;13: e0007323. doi:10.1371/journal.pntd.0007323

27. Ercumen A, Mertens A, Arnold BF, Benjamin-Chung J, Hubbard AE, Ahmed MA, et al. Effects of Single and Combined Water, Sanitation and Handwashing Interventions on Fecal Contamination in the Domestic Environment: A Cluster-Randomized Controlled Trial in Rural Bangladesh. Environ Sci Technol. 2018;52: 12078–12088. doi:10.1021/acs.est.8b05153

28. Rose G. Sick individuals and sick populations. International Journal of Epidemiology. 2001;30: 427–432. doi:10.1093/ije/30.3.427

29. VanderWeele TJ, Luedtke AR, van der Laan MJ, Kessler RC. Selecting Optimal Subgroups for Treatment Using Many Covariates. Epidemiology. 2019;30: 334–341. doi:10.1097/EDE.0000000000000991

30. Bilkey GA, Burns BL, Coles EP, Mahede T, Baynam G, Nowak KJ. Optimizing Precision Medicine for Public Health. Front Public Health. 2019;7: 42–42. doi:10.3389/fpubh.2019.00042

31. Levy J, van der Laan M, Hubbard A, Pirracchio R. A fundamental measure of treatment effect heterogeneity. Journal of Causal Inference. 2021;9: 83–108. doi:10.1515/jci-2019-0003

32. Gruber S, Van Der Laan MJ. Targeted maximum likelihood estimation: A gentle introduction. 2009.

33. van der Laan MJ, Gruber S. Collaborative double robust targeted maximum likelihood estimation. Int J Biostat. 2010;6: Article 17. doi:10.2202/1557-4679.1181

34. van der Laan M, Coyle J, Hejazi N, Malenica I, Phillips R, Hubbard A. Targeted Learning in R: Causal Data Science with the tlverse Software Ecosystem. 2023. Available: https://tlverse.org/tlverse-handbook/optimal-individualized-treatment-regimes.html

35. Murphy SA. Optimal dynamic treatment regimes: *Dynamic Treatment Regimes*. Journal of the Royal Statistical Society: Series B (Statistical Methodology). 2003;65: 331–355. doi:10.1111/1467-9868.00389

36. Zhao Y, Zeng D, Rush AJ, Kosorok MR. Estimating Individualized Treatment Rules Using Outcome Weighted Learning. Journal of the American Statistical Association. 2012;107: 1106–1118. doi:10.1080/01621459.2012.695674

37. Robins J, Rotnitzky A. Discussion of “Dynamic treatment regimes: Technical challenges and applications.” Electron J Statist. 2014;8. doi:10.1214/14-EJS908

38. Zhang B, A. Tsiatis A, Davidian M, Zhang M, Laber E. Estimating optimal treatment regimes from a classification perspective: Estimating optimal treatment regimes from a classification perspective. Stat. 2016;5: 278–278. doi:10.1002/sta4.124

39. Chakraborty B, Moodie EEM. Statistical Methods for Dynamic Treatment Regimes. New York, NY: Springer New York; 2013. doi:10.1007/978-1-4614-7428-9

40. Robins J. A new approach to causal inference in mortality studies with a sustained exposure period—application to control of the healthy worker survivor effect. Mathematical Modelling. 1986;7: 1393–1512. doi:10.1016/0270-0255(86)90088-6

41. Arnold BF, Null C, Luby SP, Unicomb L, Stewart CP, Dewey KG, et al. Cluster-randomised controlled trials of individual and combined water, sanitation, hygiene and nutritional interventions in rural Bangladesh and Kenya: the WASH Benefits study design and rationale. BMJ Open. 2013;3: e003476–e003476. doi:10.1136/bmjopen-2013-003476

42. Curtis VA, Danquah LO, Aunger RV. Planned, motivated and habitual hygiene behaviour: an eleven country review. Health education research. 2009;24: 655–673.

43. Crane RJ, Jones KDJ, Berkley JA. Environmental enteric dysfunction: an overview. Food Nutr Bull. 2015;36: S76–S87. doi:10.1177/15648265150361S113

44. Liu J, Gratz J, Amour C, Nshama R, Walongo T, Maro A, et al. Optimization of quantitative PCR methods for enteropathogen detection. PloS one. 2016;11: e0158199.

45. Liu J, Platts-Mills JA, Juma J, Kabir F, Nkeze J, Okoi C, et al. Use of quantitative molecular diagnostic methods to identify causes of diarrhoea in children: a reanalysis of the GEMS case-control study. The Lancet. 2016;388: 1291–1301.

46. Platts-Mills JA, Liu J, Rogawski ET, Kabir F, Lertsethtakarn P, Siguas M, et al. Use of quantitative molecular diagnostic methods to assess the aetiology, burden, and clinical characteristics of diarrhoea in children in low-resource settings: a reanalysis of the MAL-ED cohort study. The Lancet Global Health. 2018;6: e1309–e1318.

47. Kosek MN. Causal Pathways from Enteropathogens to Environmental Enteropathy: Findings from the MAL-ED Birth Cohort Study. EBioMedicine. 2017;18: 109–117. doi:10.1016/j.ebiom.2017.02.024

48. Amour C, Gratz J, Mduma E, Svensen E, Rogawski ET, McGrath M, et al. Epidemiology and Impact of Campylobacter Infection in Children in 8 Low-Resource Settings: Results From the MAL-ED Study. Clin Infect Dis. 2016;63: 1171–1179. doi:10.1093/cid/ciw542

49. Stephensen CB. Burden of Infection on Growth Failure. The Journal of Nutrition. 1999;129: 534S–538S. doi:10.1093/jn/129.2.534S

50. Cogill B. Anthropometric Indicators Measurement Guide. Food and Nutrition Technical Assistance Project. 2003.

51. de Onis M, Onyango AW, Van den Broeck J, Chumlea WC, Martorell R. Measurement and standardization protocols for anthropometry used in the construction of a new international growth reference. Food Nutr Bull. 2004;25: S27–36. doi:10.1177/15648265040251S104

52. van der Laan MJ, Polley EC, Hubbard AE. Super learner. Stat Appl Genet Mol Biol. 2007;6: Article25. doi:10.2202/1544-6115.1309

53. Phillips RV, van der Laan MJ, Lee H, Gruber S. Practical considerations for specifying a super learner. International Journal of Epidemiology. 2023; dyad023. doi:10.1093/ije/dyad023

54. Ivana Malenica, Coyle J, Van Der Laan M. tmle3mopttx. Available: https://github.com/tlverse/tmle3mopttx

55. Van der Laan MJ, Rose S. Targeted learning in data science: causal inference for complex longitudinal studies. New York, NY: Springer Berlin Heidelberg; 2017.

56. van der Laan MJ, Rose S. Targeted learning: causal inference for observational and experimental data. New York: Springer; 2011.

57. Schuler MS, Rose S. Targeted Maximum Likelihood Estimation for Causal Inference in Observational Studies. Am J Epidemiol. 2017;185: 65–73. doi:10.1093/aje/kww165

58. Glynn AN, Quinn KM. An Introduction to the Augmented Inverse Propensity Weighted Estimator. Political Analysis. 2017/01/04 ed. 2010;18: 36–56. doi:10.1093/pan/mpp036

59. Kuhn M. The caret Package. 2019. Available: https://topepo.github.io/caret/

60. Fitzgerald C. Campylobacter. Clin Lab Med. 2015;35: 289–298. doi:10.1016/j.cll.2015.03.001

61. Rogawski ET, Liu J, Platts-Mills JA, Kabir F, Lertsethtakarn P, Siguas M, et al. Use of quantitative molecular diagnostic methods to investigate the effect of enteropathogen infections on linear growth in children in low-resource settings: longitudinal analysis of results from the MAL-ED cohort study. The Lancet Global Health. 2018;6: e1319–e1328.

62. Penakalapati G, Swarthout J, Delahoy MJ, McAliley L, Wodnik B, Levy K, et al. Exposure to Animal Feces and Human Health: A Systematic Review and Proposed Research Priorities. Environ Sci Technol. 2017;51: 11537–11552. doi:10.1021/acs.est.7b02811

63. Butzin-Dozier Z, Waters WF, Baca M, Vinueza RL, Saraiva-Garcia C, Graham J. Assessing Upstream Determinants of Antibiotic Use in Small-Scale Food Animal Production through a Simulated Client Method. Antibiotics. 2021;10. doi:10.3390/antibiotics10010002

64. Young KT, Davis LM, DiRita VJ. Campylobacter jejuni: molecular biology and pathogenesis. Nature Reviews Microbiology. 2007;5: 665–679.

65. Black RE, Levine MM, Clements ML, Hughes TP, Blaser MJ. Experimental Campylobacter jejuni infection in humans. Journal of infectious diseases. 1988;157: 472– 479.

66. Reti KL, Tymensen LD, Davis SP, Amrein MW, Buret AG. Campylobacter jejuni increases flagellar expression and adhesion of noninvasive Escherichia coli: effects on enterocytic Toll-like receptor 4 and CXCL-8 expression. Infection and immunity. 2015;83: 4571–4581.

67. Masanta WO, Heimesaat MM, Bereswill S, Tareen AM, Lugert R, Groß U, et al. Modification of intestinal microbiota and its consequences for innate immune response in the pathogenesis of campylobacteriosis. Clinical and Developmental Immunology. 2013;2013.

68. Kalischuk LD, Leggett F, Inglis GD. Campylobacter jejuni induces transcytosis of commensal bacteria across the intestinal epithelium through M-like cells. Gut Pathog 2010; 2: 14; PMID: 21040540. 2010.

69. Riddle MS, Gutierrez RL, Verdu EF, Porter CK. The chronic gastrointestinal consequences associated with Campylobacter. Current gastroenterology reports. 2012;14: 395–405.

70. Wessells KR, Arnold CD, Stewart CP, Prado EL, Abbeddou S, Adu-Afarwuah S, et al. Characteristics that modify the effect of small-quantity lipid-based nutrient supplementation on child anemia and micronutrient status: an individual participant data meta-analysis of randomized controlled trials. Am J Clin Nutr. 2021;114: 68S-94S. doi:10.1093/ajcn/nqab276

71. McCormick BJJ, Lee GO, Seidman JC, Haque R, Mondal D, Quetz J, et al. Dynamics and Trends in Fecal Biomarkers of Gut Function in Children from 1-24 Months in the MAL-ED Study. Am J Trop Med Hyg. 2017;96: 465–472. doi:10.4269/ajtmh.16-0496

72. Grant SS, Hung DT. Persistent bacterial infections, antibiotic tolerance, and the oxidative stress response. Virulence. 2013;4: 273–283. doi:10.4161/viru.23987

73. Gomez JE, McKinney JD. M. tuberculosis persistence, latency, and drug tolerance. Tuberculosis. 2004;84: 29–44.

74. Monack DM, Mueller A, Falkow S. Persistent bacterial infections: the interface of the pathogen and the host immune system. Nature Reviews Microbiology. 2004;2: 747–765.

75. McCormick BJJ, Murray-Kolb LE, Lee GO, Schulze KJ, Ross AC, Bauck A, et al. Intestinal permeability and inflammation mediate the association between nutrient density of complementary foods and biochemical measures of micronutrient status in young children: results from the MAL-ED study. Am J Clin Nutr. 2019;110: 1015–1025. doi:10.1093/ajcn/nqz151

