## Supplemental Materials for "Treatment Heterogeneity of Water, Sanitation, Hygiene, and Nutrition Interventions on Child Growth by Environmental Enteric Dysfunction and Pathogen Status for Young Children in Bangladesh"

**Supplemental Material 1. Study interventions.**

We included four study arms: control, combined water treatment, sanitation, and handwashing (WSH), nutrition, and nutrition plus WSH (N+WSH). The control arm was passive, including no visit by a health promoter. The water treatment involved provision of chlorine tablets (Aquatabs; NaDCC) and a safe storage vessel to treat and store drinking water. Sanitation involved upgrading latrines to double pit latrines for all households in study compounds, providing child potties, and sani-scoops to remove feces from households and compounds. The handwashing intervention involved providing handwashing stations near latrines and kitchens, which included soapy water bottles and detergent soap. The nutrition intervention involved the provision of a small-quantity lipid-based nutrient supplement and age-appropriate recommendations on maternal nutrition and child feeding.

**Supplemental Material 2. Inclusion criteria.**

We included households if a resident was a pregnant mother in her first or second trimester, the household was located in rural area that was not fully submerged during monsoon season and did not have water, sanitation, hygiene, or nutrition programs ongoing or planned in the next two years.

We excluded households whose residents had plans to move during the following year, did not own their home, or drew water from a source with high iron content or high arsenic content.

We included child participants who were born to enrolled mothers meeting household inclusion criteria within six months of the baseline survey. We excluded children if their growth score fell outside of the WHO plausible range [1].

**Supplemental Material 3**. **Adjustment covariates.**

Enrollment characteristics: child sex (male or female), child birth order (first born, second born or greater), maternal age (years), maternal height (cm.) maternal education level (no education, primary, or secondary/greater), household food insecurity (4-level HFIAS categories), number of children in the household less than 18 years old, total number of individuals living in the compound (group of nearby houses), household's distance to primary drinking water source (in minutes), household construction materials (floor, walls, and roof), and an asset-based household wealth index, calculated from the first principal component of a principal components analysis of household assets (electricity, wardrobe, table, chair or bench, khat, chouki, working radio, working black/white or color television, refrigerator, bicycle, motorcycle, sewing machine, mobile phone, land phone, number of cows, number of goats, number of chickens).

Time-varying characteristics: child age (days) at assessment and month of data collection.

**Supplemental Material 4. Laboratory methods.**

*EED Biomarkers*

Study staff followed enzyme-linked immunosorbent assay kit protocols for fecal alpha-1-antitrypsin, myeloperoxidase, and REG1B.

*Pathogens*

The child's primary caregiver collected the fecal sample, it was placed on cold chain at median time 155 minutes (interquartile range, [IQR], 80–529), and then transported on dry ice to the laboratory where it was stored at -80 degrees Celsius [2]. We extracted DNA and RNA using QIAamp Fast DNA Stool Mini kit (Qiagen, Venlo, The Netherlands) as well as spike-ins of two extrinsic controls which aimed to assess efficiency of extraction and amplification [3]. We assessed enteropathogens at icddr,b using quantitative polymerase chain reaction (PCR) via TaqMan array card [3,4]. We quantified pathogens using quantification cycle, where one unit corresponded to twice the pathogen quantity and there was an analytical limit of detection at quantification cycle 35 [5]. These quantities were normalized based on the efficiency of per-sample extraction/amplification.

**Supplemental Material 5. Pathogens**

We assessed the relative concentration of the following pathogens: *Campylobacter jejuni/coli*, enteroaggregative *Escherichia coli* (EAEC; pathotypes *aaiC, or aatA, or bot)*), any enterotoxigenic *Escherichia coli* (ETEC; pathotypes *LT*, *STh*, *STp*), atypical enteropathogenic *Escherichia coli* (aEPEC; pathotypes (*eae* without *bfpA*, *stx1*, and *stx2*), typical enteropathogenic *Escherichia coli* (EPEC; pathotypes *bfpA* and *eae)*, *Campylobacter* spp., *E. coli* with heat-stable toxin, Shiga toxin–producing *E. coli*, *Shigella*/enteroinvasive *E. coli*, *Ancyclostoma, Necator, E. bieneusi, E.intestinalis*, *E.histolytica*, *Entamoeba* spp., *Giardia, Cryptosporidium, Salmonella, H. nana, Schistosoma, B. fragilis, H. pylori,* rotavirus, *Ascaris, Trichurism, Cyclospora, Isospora, Cryptosporidium hominis, Cryptosporidium parvum, Strongyloides, Blastocystis, V. cholerae, M. tuberculosis, C. difficile, Plesiomonas, Aeromonas*, astrovirus norovirus GI/GII, sapovirus, and adenovirus 40/41 [2].

**Supplemental Material 6. Covariate, EED biomarker, and pathogen exclusion**

We excluded the following variables due to missingness greater than 30%, all of which were assessed at median 3 months of age: lactulose concentration, mannitol concentration, myeloperoxidase, alpha-1-antitrypsin, and age and month of assessment for stool and urine tests.

We excluded the following covariates due to near zero variance: maternal education, household food security, distance to water source, household floor material, enterotoxigenic *E. coli* with heat-labile toxin, enterotoxigenic *E. coli* with heat-stable toxin, typical enteropathogenic *E. coli*, Shiga toxin–producing *E. coli* (STEC), *Shigella*/enteroinvasive *E. coli*, *Ancyclostoma*, *Necator*, *E. bieneusi, E.intestinalis*, *E.histolytica*, *Entamoeba* spp., *Giardia*, *Cryptosporidium, Salmonella, H. nana, Schistosoma, B. fragilis*, *H. pylori*, rotavirus, *Ascaris, Trichurism, Cyclospora, Isospora*, *Cryptosporidium hominis, Cryptosporidium parvum, Strongyloides, Blastocystis, V. cholerae, M. tuberculosis*, *C.difficile*, *Plesiomonas, Aeromonas*, and astrovirus.

**Supplemental Material 7. Correlation of individual N+WSH conditional treatment effect and myeloperoxidase concentration**

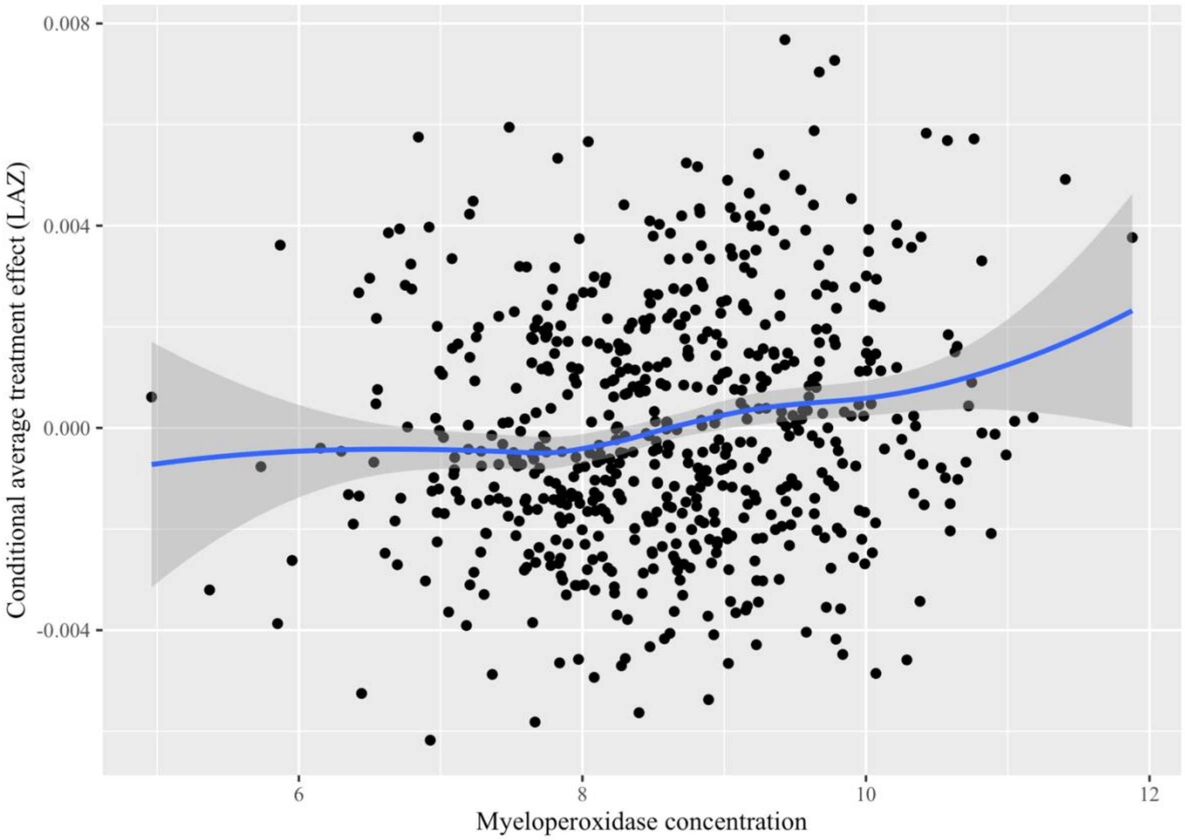


N+WSH, combined nutrition, water, sanitation, and hygiene intervention; LAZ, length-for-age z-score

**Supplemental Material 8. Correlation of individual N+WSH conditional treatment effect and *Campylobacter* concentration**

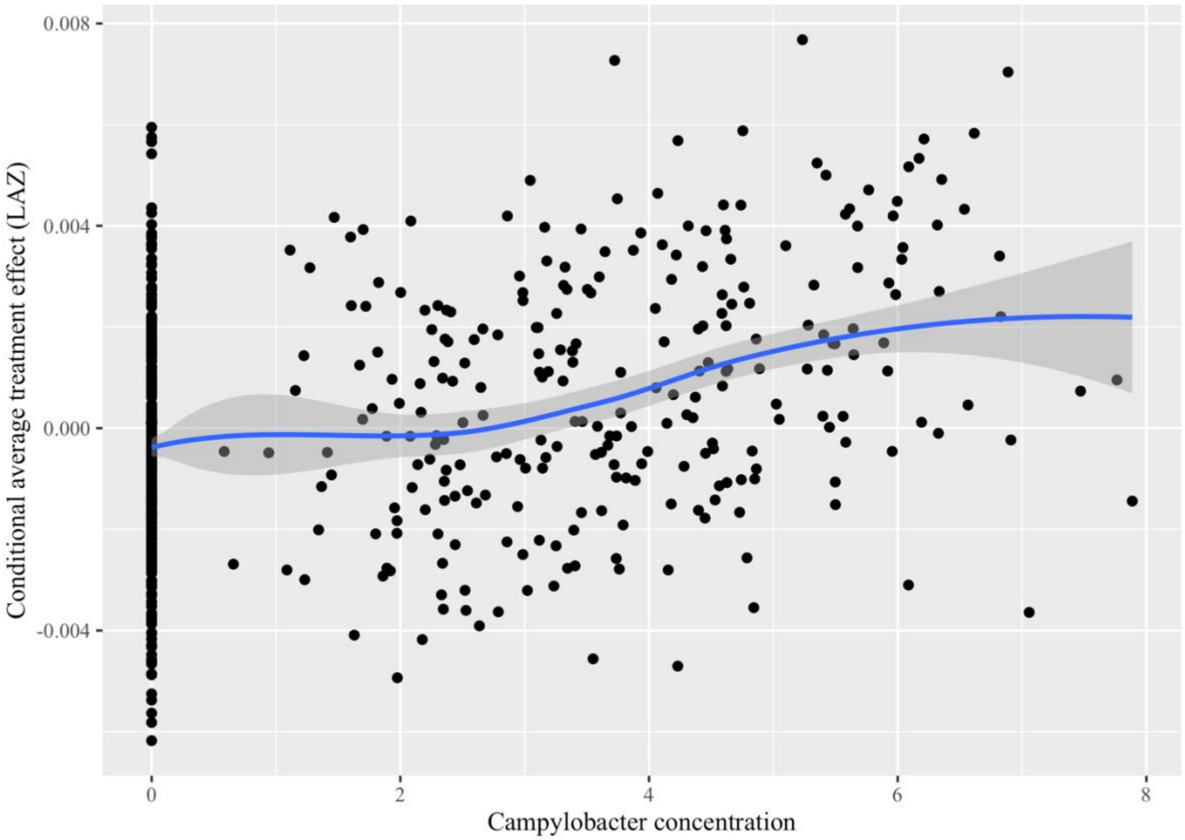


N+WSH, combined nutrition, water, sanitation, and hygiene intervention; LAZ, length-for-age z-score

**Supplemental Material 9. Correlation of individual WSH conditional treatment effect and myeloperoxidase concentration**

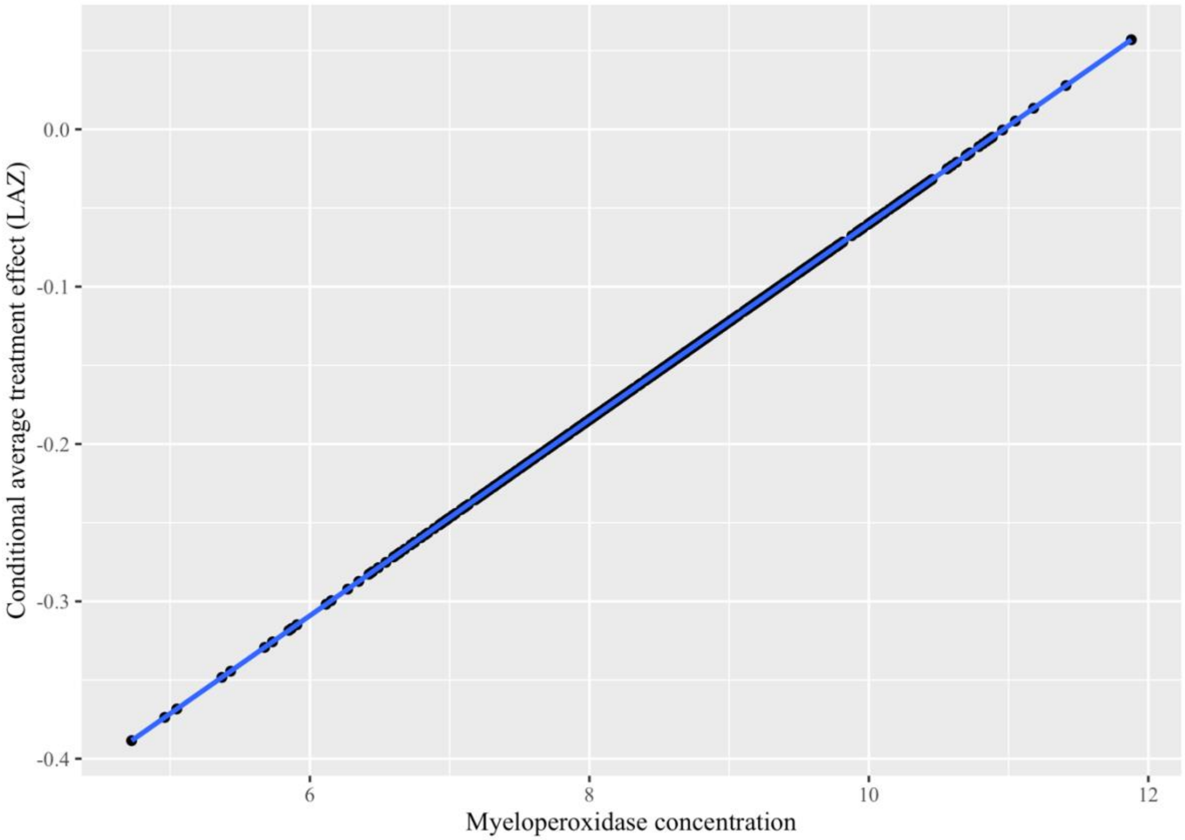


WSH, water, sanitation, and hygiene intervention; LAZ, length-for-age z-score

**Supplemental Material 10. Correlation of individual WSH conditional treatment effect and *Campylobacter* concentration**

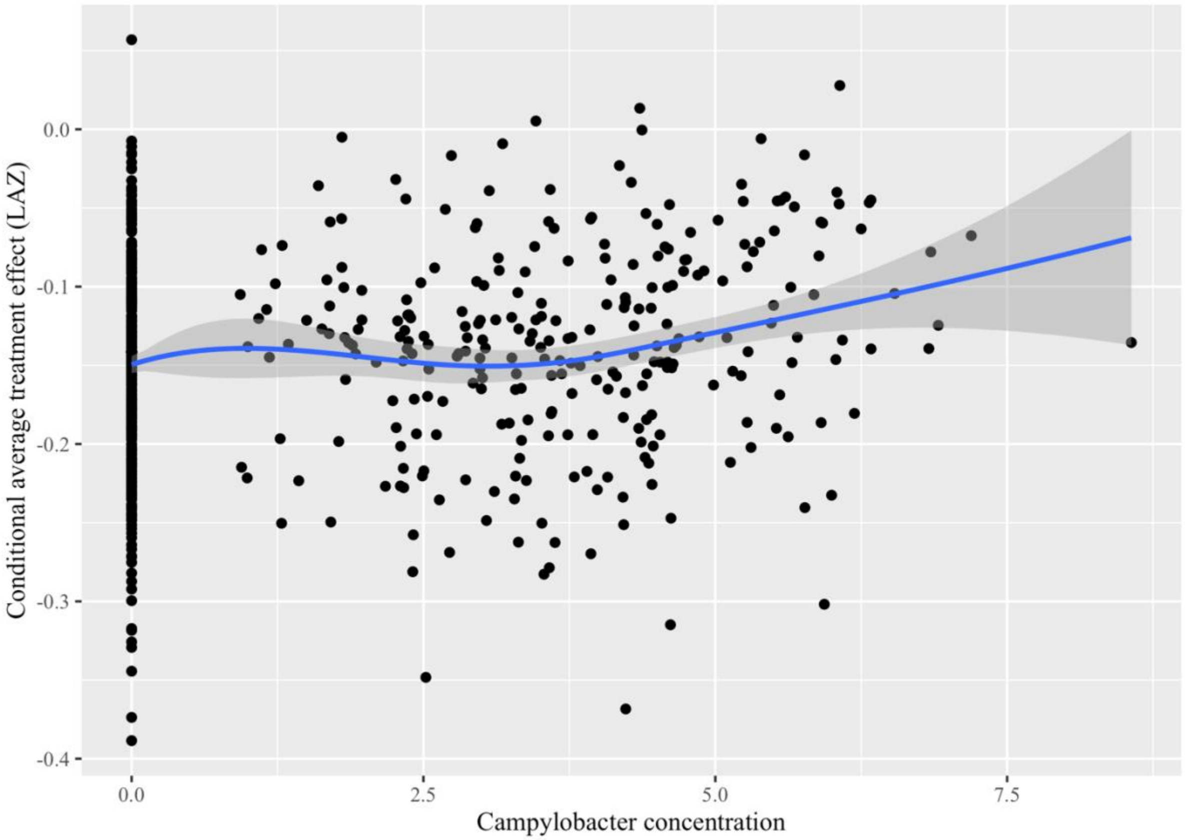


WSH, water, sanitation, and hygiene intervention; LAZ, length-for-age z-score

**Supplemental Material 11. Correlation of individual nutrition conditional treatment effect and myeloperoxidase concentration**

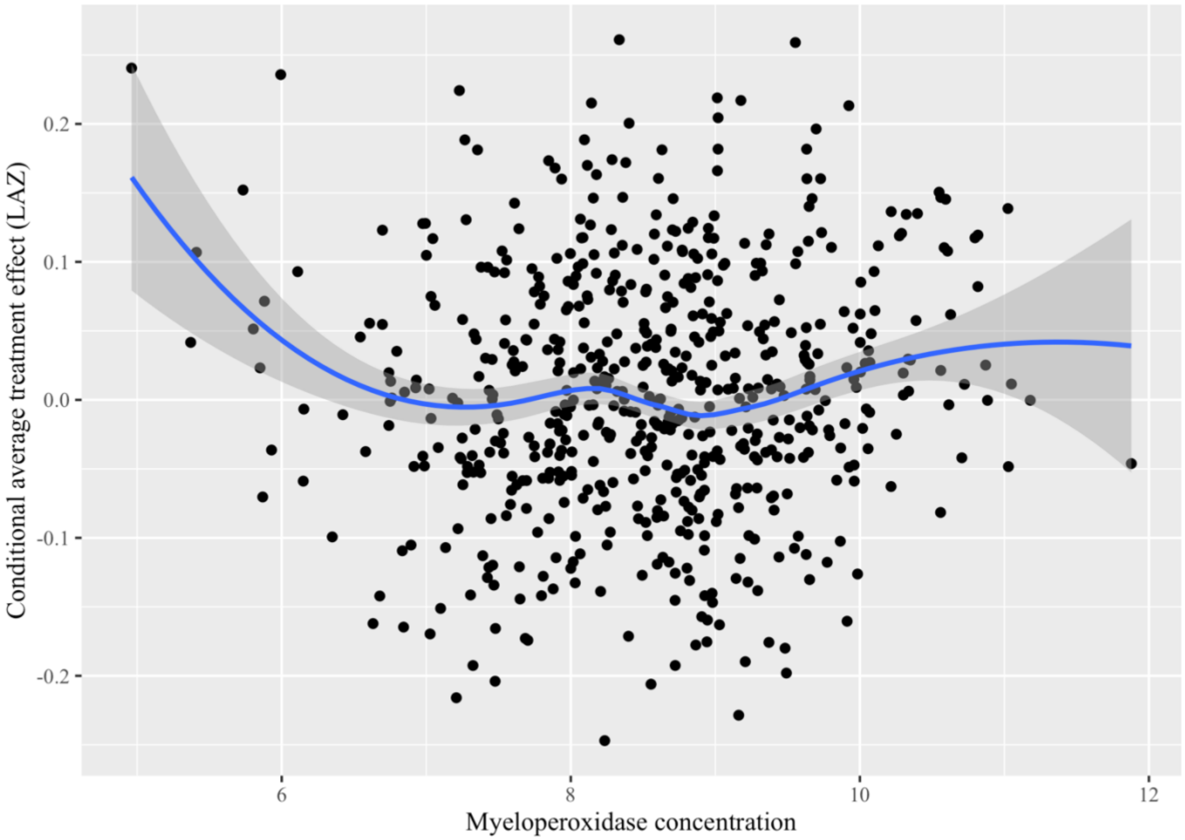


LAZ, length-for-age z-score

**Supplemental Material 12. Correlation of individual nutrition conditional treatment effect and *Campylobacter* concentration**

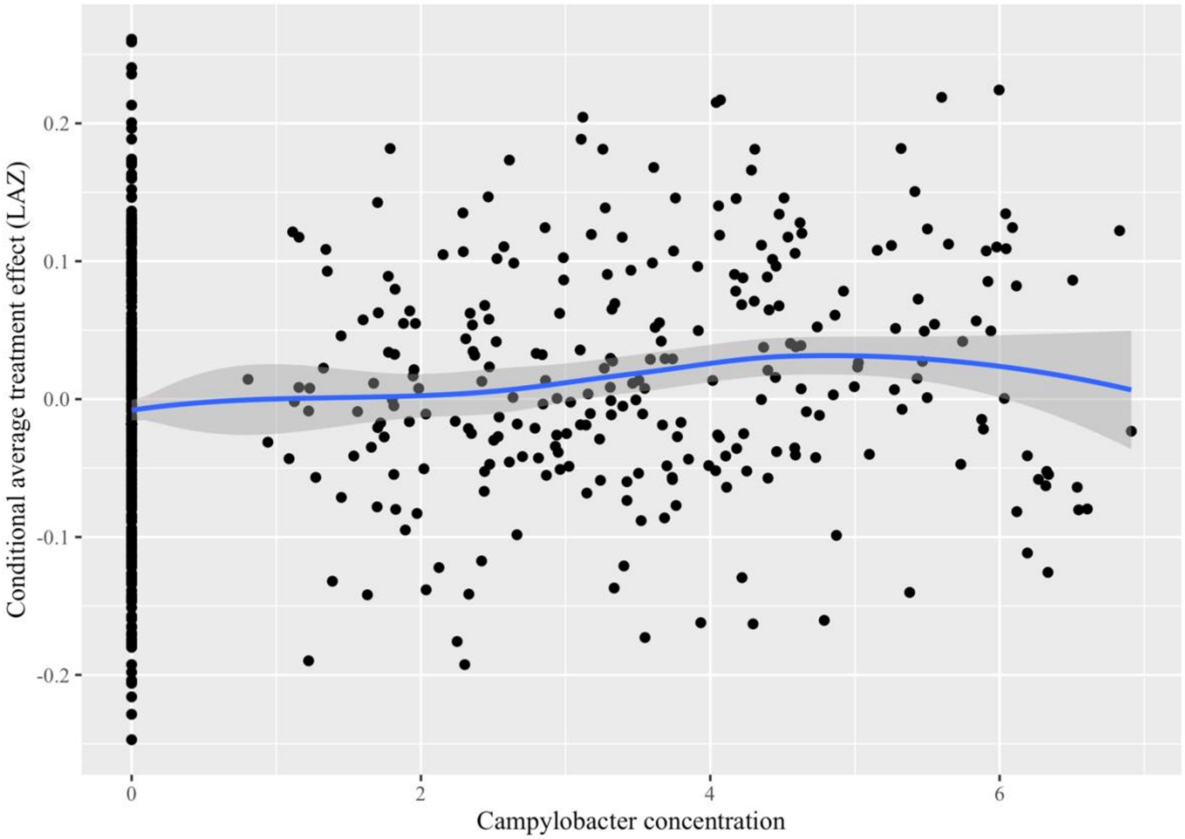


LAZ, length-for-age z-score

**Supplemental Figure 11: Enrollment flowchart.**

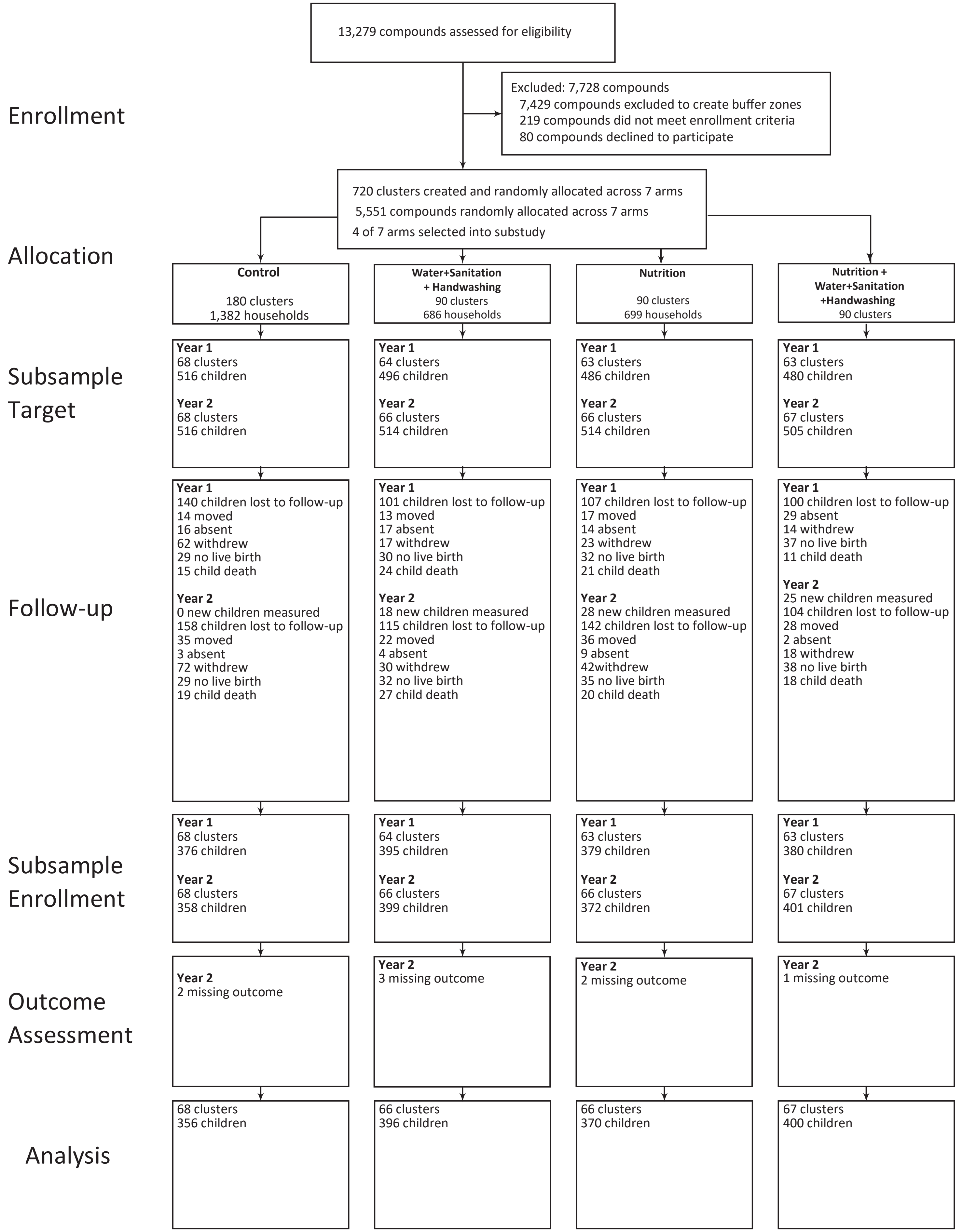

REFERENCES


1. de Onis M, Onyango AW, Van den Broeck J, Chumlea WC, Martorell R. Measurement and standardization protocols for anthropometry used in the construction of a new international growth reference. Food Nutr Bull. 2004;25: S27-36. doi:10.1177/15648265040251S104

2. Grembi JA, Lin A, Karim MA, Islam MO, Miah R, Arnold BF, et al. Effect of water, sanitation, handwashing and nutrition interventions on enteropathogens in children 14 months old: a cluster-randomized controlled trial in  rural Bangladesh. J Infect Dis. 2020. doi:10.1093/infdis/jiaa549

3. Liu J, Gratz J, Amour C, Nshama R, Walongo T, Maro A, et al. Optimization of quantitative PCR methods for enteropathogen detection. PloS one. 2016;11: e0158199.

4. Liu J, Platts-Mills JA, Juma J, Kabir F, Nkeze J, Okoi C, et al. Use of quantitative molecular diagnostic methods to identify causes of diarrhoea in children: a reanalysis of the GEMS case-control study. The Lancet. 2016;388: 1291–1301.

5. Platts-Mills JA, Liu J, Rogawski ET, Kabir F, Lertsethtakarn P, Siguas M, et al. Use of quantitative molecular diagnostic methods to assess the aetiology, burden, and clinical characteristics of diarrhoea in children in low-resource settings: a reanalysis of the MAL-ED cohort study. The Lancet Global Health. 2018;6: e1309–e1318.
